# “They recognize me as a doctor”: evaluation of an HIV peer mobilisation training programme to promote oral HIV self-testing and referral for acute HIV infection screening among gay and bisexual men and transgender women in coastal Kenya

**DOI:** 10.1101/2025.04.08.25324345

**Authors:** Shaun Palmer, Maartje Dijkstra, Alex Kigoro, Khamisi Mohamed, Nana Mukuria, Shally Mahmoud, Evanson Gichuru, Elise M van der Else, Eduard J. Sanders

## Abstract

**Background:** Targeted peer mobilisation can improve access to HIV testing and care and may impact onward HIV transmission. We describe the development and implementation of a peer mobilisation training programme for oral HIV self-testing (OST) and referral for acute HIV infection testing among gay and bisexual men (GBMSM) and transgender women (TGW) in coastal Kenya.

**Methods:** The training programme covered five modules: 1) safe sex, 2) OST, 3) acute HIV infection (AHI), 4) HIV partner notification services, and 5) mobilisation skills. Mobilisers attended two training sessions and weekly meetings between March and June 2019. Mobilisers offered OST to GBMSM and TGW peers and extended an AHI referral card for point-of-care HIV-RNA testing when peers reported AHI symptoms. Two focus group discussions with 18 mobilisers and 15 in-depth interviews with mobilised clients who were newly HIV diagnosed were conducted to assess which components of the training programme were most helpful in mobilising clients for HIV testing.

**Results:** Mobilisers felt empowered through the training programme, which enhanced their mobilisation skills across two areas: (1) networking skills and (2) client empowerment. Facilitators for HIV testing were confidentiality of the OST, presence of STI symptoms, and building trust between mobilisers and clients. Mobilisers and clients reported challenges as: (1) misconceptions regarding OST and symptoms of AHI, (2) logistical and financial issues, and (3) stigma and security concerns.

**Discussion:** Our training programme facilitated peer mobilisers to extend OSTs among GBMSM and TGW in coastal Kenya while it was more difficult to refer clients directly for AHI testing. Mobilisers felt empowered through enhanced mobilisation skills which helped them to mobilise clients for HIV testing. A targeted training programme was helpful in mobilising peers to take up HIV testing.

## Introduction

Gay and bisexual men and transgender women who have sex with men (GBMSM and TGW^1^) represent key and vulnerable populations for HIV acquisition in Kenya. A 2019 study in coastal Kenya demonstrated an incidence of 5.1 (95% confidence interval [CI], 2.6–9.8) per 100 person-years (PY) among GBMSM and 20.6 (95% CI, 6.6–63.8) per 100 PY among TGW (1). Tailored approaches to engage people in HIV testing, by way of peer mobilisation, can help prevent HIV transmission among GBMSM and TGW if newly diagnosed participants can be linked to care and HIV negative participants initiated on PrEP (2–7). Targeted testing approaches such as oral self-testing (OST) can reduce access barriers to testing among key and vulnerable populations (5,8). While OST can diagnose chronic HIV infection, it will miss cases of acute HIV infection (AHI). Individuals presenting with symptoms of AHI can be referred for AHI testing if point-of-care RNA testing is available. It is estimated that 30-50% of onward transmission events occur during AHI (9–12).

Same-sex sexual behaviour is criminalised in Kenya, based on British colonial law, and stigma against GBMSM and TGW persists in communities and during care-seeking at health facilities (13–15). Peer mobilisation represents an effective way to target GBMSM and TGW who have been unsuccessfully engaged for HIV testing (4–7), with peer mobilisers (*henceforth: mobilisers*) viewed as credible sources of sexual health information by their peers (2,3). GBMSM and TGW in sub-Saharan Africa are also often part of large and active social and sexual networks (16). Peer mobilisation can be delivered systematically and discretely within these networks in a social climate where more overt strategies, such as targeted publicity campaigns, are not possible (6,7,16,17).

Extensive input from the local Lesbian, Gay, Bisexual, Transgender, Queer, Intersex (LGBTI) community is a prerequisite to reflect the local socio-cultural context (9,17).

Knowledge about AHI among GBMSM and TGW is low, thus health education campaigns should include information on this topic (9,18–26). AHI occurs during the first weeks after infection, characterised by a peak in viral load during which anti-HIV-1 antibodies are undetectable but HIV-1 RNA or p24 antigen are present (10,27,28). While detecting AHI is rare, targeting GBMSM and TGW for HIV testing must include information about symptoms of AHI and AHI referral pathways (29).

Our research team earlier described the feasibility and effectiveness of a peer mobilisation-led strategy in the delivery of HIV partner notification services, OST and AHI testing referral, and linkage to ART and PrEP services in coastal Kenya (29–31). As part of this parent study, we developed and implemented a training programme for GBMSM and TGW peer mobilisers about sexual health, and OST and AHI testing mobilisation. Here, we assessed the experience with this training programme and peer mobilisation from the perspective of the peer mobilisers and their mobilised clients. We identified facilitators for and challenges with peer mobilisation for OST and referral for AHI testing.

## Methods

### Study setting and population

Between March and June 2019, we delivered a three-month peer mobilisation training programme on sexual health, OST, and AHI testing referral to GBMSM and TGW, in the context of the parent study on peer mobilisation-led HIV testing (32). The study took place at two Kenyan Medical Research Institute (KEMRI) sites in Kilifi county: 1) Malindi Sub-County Hospital and 2) KEMRI-–Wellcome Trust Research Programme (KEMRI-KWTRP) research clinic Mtwapa. These clinics provided HIV testing and counselling services (HTC) and routinely engaged GBMSM and TGW in research (33,34). Mobilisers were purposively selected by clinic staff due to participation in previous studies and being representative of their GBMSM and TGW peers. Mobilisers were 18 years or older and cisgender males or transgender women (TGW) who have sex with men. The majority were living with HIV.

### Parent study overview

The parent study investigated the feasibility and effectiveness of a peer-mobilisation self-testing strategy, combined with AHI testing and HIV partner notification services (HPN) for GBMSM and TGW (32). Detailed study procedures of the parent study can be found elsewhere (32). In short: each week, six to 10 mobilisers at each study site were instructed to recruit five GBMSM and TGW peers for HIV testing via their peer network, including via hot spots and social media. Peers were behaviourally vulnerable to HIV infection, defined as aged 18-24 years old and engaging in one or more of the following behaviours: group sex, multiple sexual partners, and condomless receptive anal intercourse, in accordance with a risk score previously developed by our study team (33).

Mobilised peers (*hereafter: clients*) were eligible to receive three types of HIV test. Mobilisers first provided OSTs to their clients. Clients were encouraged to use the OST at home or at a study clinic. All clients were then invited to report to a study clinic for study enrolment, which included confirmation testing at the clinic via HIV rapid antibody testing and point-of-care HIV RNA testing via GeneXpert™. Mobilisers were also instructed to identify clients with AHI symptoms and to immediately refer them to the study clinic for a direct point-of-care RNA test by giving them an AHI referral card to present at the clinic. AHI symptoms, referred to as malaria-like symptoms, included fatigue, fever, sore throat, body pain, and diarrhoea, and genital ulcers.(30,35).

Clients newly diagnosed with HIV were invited to enrol as participants to undergo HPN as part of the parent study and were reimbursed 350 Kenyan Shillings (KSh). Mobilisers were reimbursed 1500 KSh (∼USD 15) weekly once five peers had attended the clinic for confirmation testing. They additionally received 500 KSh (∼USD 5) weekly for travel reimbursement. There was no additional target or reimbursement for AHI referral as it was unclear how many symptomatic peers they would meet during the recruitment period.

### Patient Consent Statement

The KEMRI Scientific Ethical Review Unit approved the study (135/3747). All participants provided written informed consent prior to enrollment in the parent study following peer mobilisation.

### Training programme

The training programme included two training sessions, weekly meetings during the three month study period, and communication materials (figure 1.). The programme covered five modules: 1) safe sex, 2) OST, 3) AHI, 4) HPN, and 5) peer mobilisation skills (29,30,33). The training programme and communication materials were designed collaboratively with LGBTI study staff, representatives from two local LGBTI organisations, and mobilisers. We sought feedback on the development, comprehension, and relevance of the content and translations at preparatory meetings. All materials were developed in English and Kiswahili. Each mobiliser received print and digital versions of the communication materials to share among clients. A WhatsApp group was also created for mobilisers and study staff to share ideas and resources and facilitate a dynamic learning process.

**Figure 1.**
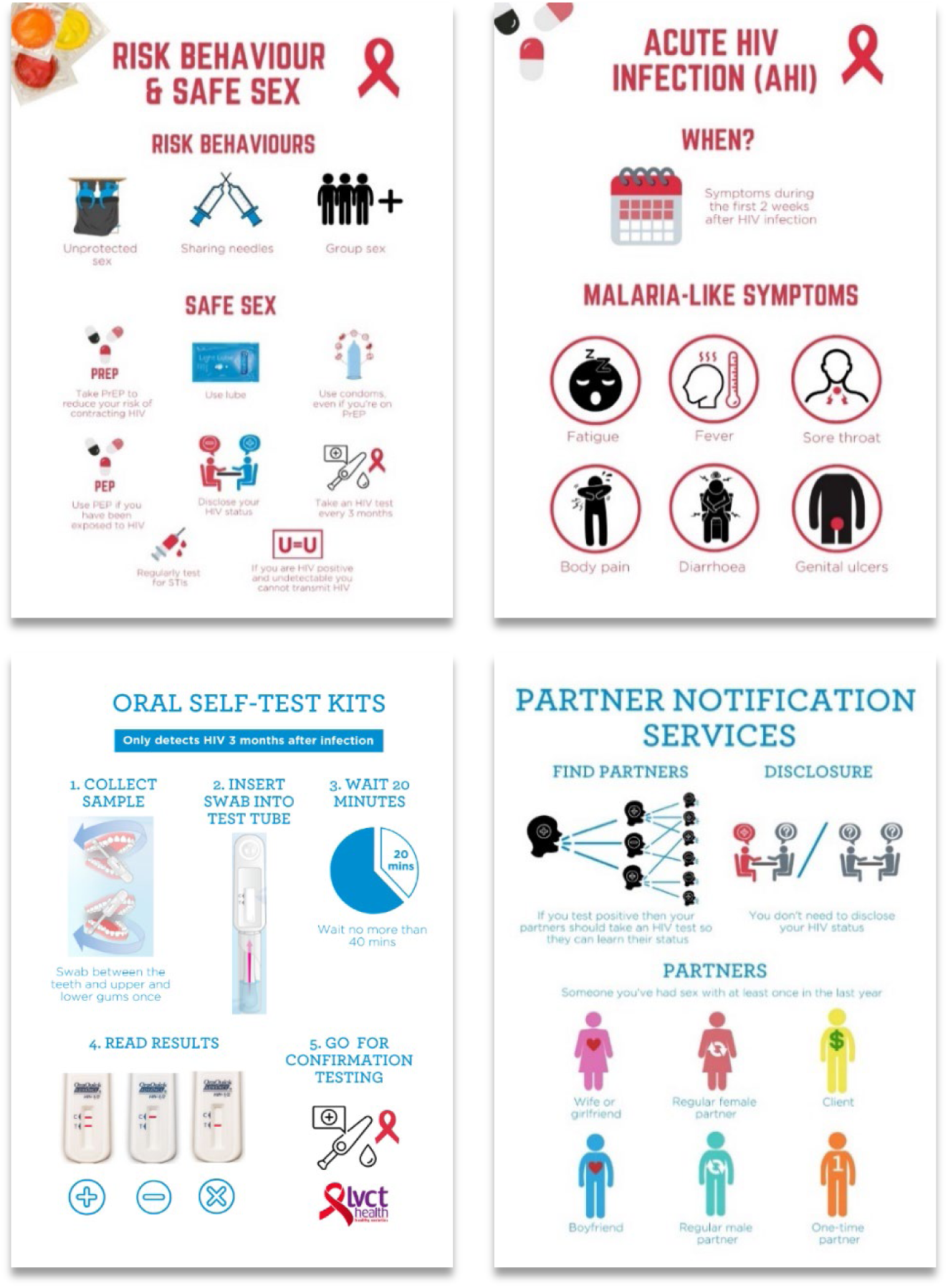
Example of pages from the English communication materials.

#### Training and weekly meetings

At the programme’s inception, 16 mobilisers attended a full-day training session facilitated by LGBTI KEMRI clinic staff covering the five modules. The session included role plays to practice peer mobilisation skills. We held a second training session six weeks later, attended by 11 new mobilisers and by the 16 existing mobilisers to review the modules. During weekly meetings, experiences with mobilisation were discussed, communication materials were reviewed, and mobilisers received new OSTs and AHI referral cards. The training sessions and weekly meetings were led by two researchers and one or two HIV counsellors or outreach workers with extensive experience working with GBMSM and TGW. The study team provided daily supervision to the mobilisers, in person and via telephone, to review study procedures and address issues such as scheduling conflicts.

### Data collection and analysis

We qualitatively assessed how the training programme was received through two focus group discussions (FGD) with mobilisers and in-depth interviews with clients who were newly diagnosed with HIV through peer mobilisation. Participants of the FGDs and in-depth interviews (IDI) were selected via convenience sampling based on availability and willingness to participate. FGDs lasted approximately two and a half hours, interviews up to one hour. We explored mobilisers’ and clients’ experience with the mobilisation process. FGDs explored the challenges mobilisers faced with mobilising clients, how the training programme influenced their role and how it could be improved, and how we can better identify AHI among GBMSM and TGW. In-depth interviews explored motivations for HIV testing, how clients experienced the information and support received from mobilisers, and recommendations for future mobilisation strategies. Participants were reimbursed 500 KSh (∼USD 5). The FGDs and IDIs were moderated, transcribed, and translated by three native bilingual (English and Kiswahili) members of the study team. Transcripts were coded using atlas.ti 8 for Windows by three members of the study team independently and inconsistencies between coders were resolved by discussion. Two researchers conducted the subsequent analysis. A thematic analysis was employed to highlight similarities and differences across the dataset while generating unanticipated insights throughout the coding and analysis (36). Extensive fieldnotes recorded during training events and weekly meetings informed the thematic analysis.

## Results

Eighteen of the 27 mobilisers who were engaged in the parent study participated in the two FGDs. Participants had a median age of 26.5 years (interquartile range [IQR]: 24-30), half were TGW, half identified as bisexual, and less than a quarter (22.2%) identified as gay (Table 1). Most participants had secondary level education or less. All had engaged in sex work in the last six months. All had experience with peer mobilisation, including mobilising with OSTs in previous studies, with a median number of years of experience of 4.5 years (IQR: 2-10).

**Table 1.** Demographic characteristics of 18 Focus group discussion participants.

|  |  | Frequency (%) |
| --- | --- | --- |
| **Gender** | Male | 9 (50.0) |
|  | Transgender woman | 9 (50.0) |
| **Sexuality** | Gay | 4 (22.2) |
|  | Bisexual | 9 (50.0) |
|  | Other | 5 (27.8) |
| **Religion** | Christian | 10 (55.6) |
|  | Muslim | 6 (33.3) |
|  | Other | 2 (11.1) |
| **Highest level of education completed** | Primary | 6 (33.3) |
|  | Secondary | 7 (38.9) |
|  | Tertiary | 4 (22.2) |
|  | Prefer not to say | 1 (5.6) |
| **Received payment for sex in the last six months** | Yes | 18 (100.0) |
| **Experience as peer mobiliser** | Yes | 18 (100) |
| **Experience mobilising with OSTs** | Yes | 18 (100) |
|  |  | **Median (IQR)** |
| **Age (years)** |  | 26.5 (24-30) |
| **Median number of years of experience as peer educator/mobiliser** |  | 4.5 (2-10) |

Out of the 23 clients who were mobilised through peer mobilisation in the parent study and identified to be living with HIV, fifteen participated in IDIs (two with early HIV infection, 13 with chronic HIV infection, and three were later reclassified as know positive). All clients interviewed were male and identified as bisexual. The median age was 25.5 years old (IQR: 22-29).

### Experience with the training programme

The mobilisers described the training programme as a valuable and informative process through which they developed their mobilising skills. They highlighted the interactive format of the training sessions and regular interactions with the study team during weekly meetings as key to helping them understand and remember the training content. They described feeling able to provide accurate information to their clients, including on HIV symptoms, how to use an OST, and the importance of knowing their HIV status. They further commented that the weekly meetings motivated them in their role to regularly discuss challenges and identify solutions together with their fellow mobilisers and the study team. The communication materials were also found to be useful reference materials by the mobilisers, however, a substantial number of clients reported that the communication materials were not shared with them.

> *“I think they [the training sessions and weekly meetings] were very helpful because we used to remind each other of things we might have forgotten because some of us were starting to maybe forget their roles and in those meeting[s], we help each other out.” **[Mobiliser, male, 31-35 years old]***

### Mobilisation skills

Mobilisers described how the training programme enhanced their mobilisation skills under two thematic areas: 1) networking skills, and 2) client empowerment.

#### Networking skills

Mobilisers described how both the training programme and the mobilisation process improved their networking skills, including their confidence. They reported feeling empowered, noting that the training programme helped them learn how to approach clients and how to talk to them and educate them about sensitive topics in a discrete and confidential way.

> *“Personally, I was not able to talk to people one on one, but it has helped me a lot in terms of socialisation and courage wise.” [**Mobiliser, TGW, 26-30 years old**]*

Mobilisers reported that in-person mobilisation was most effective at hotspots where they had existing clients, or at parties, gyms, and local LGBTI organisations. Online forums, such as WhatsApp, Grindr, and Gay Romeo, was cited as another, although less common, route to initiate mobilisation. In these spaces, mobilisers reported being viewed as credible information sources which led to clients referring other peers for mobilisation. This was corroborated by several clients.

> *“Some approach me and want services… though I am not a doctor they recognize me as a doctor and I help them by taking them to [the clinic]” **[Mobiliser, male, 21-25 years old]***

#### Client empowerment

Both mobilisers and clients spoke about the important role of mobilisers in empowering clients to test for HIV. Mobilisers would usually contact their clients several times before their clients would agree to test. Mobilisers and clients described that this required the mobilisers demonstrate considerable patience and perseverance in order to develop a relationship and build trust with their clients. Through this, the mobilisers could also mitigate stigma and address misconceptions about HIV testing procedures.

> *“He looked at my condition and advised me to come for testing, but I was not convinced at all. In a period of two weeks he came to me again and told me it was important to come to the hospital and seek help, that’s when I gave myself courage and came along with him to this place and was tested for HIV.” **[Client, male, age not reported**]*

Clients noted that discretion and confidentiality were key to successful mobilisation. Several mobilisers and clients also shared that mobilisers would disclose their own HIV status to demonstrate the benefits of testing and build trust. Mobilisers would then typically accompany clients to the clinic and support them while testing for HIV.

> *“They were not willing to comply, so I used to tell them about my status and how I was. So they gained confidence from that and they decided to test and accepted to come here.” **[Mobiliser, TGW, 26-30 years old]***

### Facilitators and motivations for HIV testing

Mobilisers and clients reported several common facilitators and motivations for HIV testing. These included facilitators related to the specific testing approach, such as the convenience and confidentiality of the OST and a perception that the point-of-care HIV RNA test was trustworthy. They also mentioned motivations to test in general, such as health concerns or a desire to know their HIV status, including due to current or recent presentation of AHI symptoms. Several mobilisers and clients commented that the presence of genital ulcers or discharge were the reason for AHI referral.

> *“They [clients] like tell me that maybe they feel pain when they urinate and its then I start asking them questions and try to link the symptoms and inform them on the benefits of having early treatment before they become chronic and that maybe they are STIs.” **[Mobiliser, TGW, 26-30 years old**]*

### Challenges with mobilisation

Challenges with mobilisation fell under three categories: 1) misconceptions regarding OST and AHI, 2) logistical and financial issues, and 3) stigma and security concerns.

#### Misconceptions regarding OST and AHI referral

OST was a new testing approach for some clients, requiring time to inform and sensitise those that they can test for HIV orally.

> *“When I received the results, I didn’t believe, I didn’t trust the OST completely since I knew that if one is tested that blood must be drawn first. That’s why I ignored the OST results and asked the peer educator to take me with the others to [the clinic] and he agreed.” [**Client, male, 21-25 years old**]*

Mobilisers described challenges in mobilising via AHI referral as AHI symptoms and the point-of-care HIV RNA test were new and complex concepts for clients. Most clients interviewed did not recall discussing AHI with their mobiliser. Mobiliser also reported that some clients were reluctant to take a confirmatory HIV RNA test as they already had an OST result, while others were reluctant to draw blood.

Some mobilisers reported meeting no clients with AHI symptoms during the study period. Mobilisers commented that it was difficult to make AHI referrals alongside OST mobilisation due to a lack of target or reimbursement for AHI referrals. Further, mobilisers highlighted that clients did not readily associate malaria-like symptoms with AHI and often sought treatment at pharmacies or took painkillers.

> *“Firstly, you tell a client about AHI and the symptoms and that they are malaria-like, so clients don’t see the need to test because you used ‘malaria-like’ and they just take painkillers and feel a little better and they don’t bother about testing. So they are in denial and ignore that it might be AHI.” [**Mobiliser, TGW, 26-30 years old]***

#### Logistical and financial issues

Mobilisers reported that some clients would cancel appointments without sufficient notice, while others were only available during evenings or weekends when the study clinics were closed.

Mobilisers and clients reported insufficient phone credit and money for transport as a barrier to testing. Heavy rains during the rainy season (May and June) also prevented clients attending clinics.

> *“My challenge was finding the right time for my clients [peers] because most of them are working class and they are only available during the weekends and in the evening, and they sometimes suggest that the doctors personally take those services to them.” **[Mobiliser, TGW, 21-25 years old]***

#### Stigma and security concerns

Mobilisers reported experiencing stigma and threats of violence during the study period, including instances of verbal altercations and blackmail from the general community. They also received threats from clients to disclose the mobiliser’s sexual orientation. Mobilisers commented that some clients were reluctant to associate with them or the study clinics, out of fear of being perceived as gay. One transgender mobiliser detailed how TGW are more visible and may not leave home during daylight hours out of fear for safety. Local transport hubs were deemed particularly unsafe due to experiences of harassment.

> *“My experience with trans[gender women] is that they mostly stay indoors during the day… and when people come across a trans person, they will question their gender… and they might get beaten up and so… they are afraid of what will happen to them.” **[Mobiliser, TGW, 21-25 years old]***

### Recommendations for mobilisation

Recommendations made by mobilisers fell into three categories: 1) continued involvement of mobilisers in the research outreach; 2) additional training and dissemination activities; and 3) tailored outreach to other key and vulnerable populations.

#### Continued involvement of mobilisers

The mobilisers expressed a strong desire to continue in their role, including to inform training and mobilisation procedures.

> *“We should not be forgotten because we already have peers, and some are already used to us… we will have a hard time cutting off communication with clients and they will lose morale in their regular checkups and we will not be able to make follow ups because of money.” **[Mobiliser, male, 21-25 years old]***

#### Additional training and dissemination activities

Both mobilisers and clients recommended regular mobilisation to promote HIV testing including via youth groups, hotspots, and educational community meetings. Additional training was recommended to increase the knowledge of the mobilisers, particularly concerning AHI.

> *“I’m suggesting that in forums and gatherings we should be told more about HIV, the importance of frequent testing and dangers of not knowing your status in time. If people get to know more about the self-testing kits this will help people feel the ease of getting to know their HIV status and giving them the privacy they need.” **[Client, male, 26-30 years old]***

#### Tailored outreach to other key and vulnerable populations

There were repeated calls for targeted efforts to mobilise TGW in order to meet their specific needs as they are often not engaged through outreach tailored to GBMSM. GBMSM and TGW clients in rural areas were also flagged as a key group not reached by existing efforts.

> *“What I can add on that is trans people are really something we forget to look for, so it is up to us as peer mobilizers to probe for the issues and educate them and have them over the center for their own benefits.” **[Mobiliser, male, 31-35 years old]***

## Discussion

In this study, we showed that our HIV peer mobilisation training programme empowered peer mobilisers to continuously engage their clients, which ultimately led to OST and HIV-RNA testing among GBMSM and TGW in coastal Kenya. Our study demonstrates the potential of regular engagement with training providers in developing the mobiliser’s networking skills and facilitating the continued empowerment of clients to test for HIV. Mobilisers and their clients valued the peer mobilisation approach that was delivered through this study and peer mobilisers were motivated to continue in this role in the future, including through receiving further training, with mobilisation services also providing an additional source of income for the mobilisers. These findings support the results of the parent study, where we showed that the mobilisation process was effective in identifying GBMSM with undiagnosed HIV infection and linking them to care (32). Our study corroborates recent efforts in sub-Saharan Africa that demonstrate the potential of peer mobilisation strategies in facilitating HIV testing (37–39). Key populations should be targeted for HIV testing and engagement in HIV care and prevention services, including initiation of oral daily PrEP when HIV negative. In this context, our study offers several lessons.

Peer mobilisation often required mobilisers to meet multiple times with clients to build trust before clients felt empowered to test for HIV. This dynamic support network likely contributed to successful mobilisation and case finding in a context where GBMSM and TGW are highly stigmatised and criminalised. In this regard, future research can also explore the potential benefits of collaborating with local LGBTI organisations to facilitate the uptake of HIV prevention and care services among marginalised communities in order to better understand the role that such groups play in promoting health equity and reducing disparities in HIV care.

AHI was a difficult concept for both mobilisers and clients to understand, notably concerning the symptoms and the window period. This in line with findings from a recent study, showing that AHI is poorly understood among people vulnerable to acquiring HIV in Eswatini (40). Mobilising via both OST and AHI symptom referral may have been too complex and future studies would benefit from focusing on only one aspect. Recognising symptoms of AHI at the time of PrEP initiation is important and PrEP initiation should be delayed if AHI is suspected (41,42). Moreover, conceptualising symptoms as ‘malaria-like’ may have confused peers, as some sought treatment elsewhere, which has been described previously (43). Referrals for AHI and HIV testing due the presence of STI symptoms should be further explored and is supported by findings of previous studies showing that HIV testing yield is higher among people with STI symptoms (32,44,45).

Mobilisers and clients risked exclusion and verbal and physical violence from peers and the wider community due to systemic homophobia and transphobia (13,14). This presented safety and wellbeing concerns, which required considerable support from the study team to explore challenges and solutions, in line with previous findings in Peru (4). Risk assessments and safety protocols should be in place for daily use by a study team and the local LGBTI community that explicitly mitigate and respond to risks while conducting studies with GBMSM and TGW in similar contexts.

TGW remained on the periphery of mobilisation efforts. Although half of the mobilisers in our study identified as TGW, we previously reported that only seven (1.6%) of the 429 mobilised clients reported to be TGW (32). Future research should explore HIV testing mobilisation approaches tailored to TGW peers as a distinct population, such as testing services at home, testing outside clinic hours, and online support (1,46,47), and explore strategies to mitigate stigma and security concerns.

### Limitations

There were several limitations to our study. Firstly, we used convenience sampling rather than purposive sampling for the FGDs and IDIs. Secondly, we were limited to two FGDs with mobilisers, and not able to conduct any IDIs with clients that refused mobilisation and HIV testing, or with clients who received a negative HIV test result following mobilisation. These limitations may reduce the generalisability of our results to the broader GBMSM and TGW community. While knowledge evaluation with surveys pre- and post-training can assess knowledge uptake (18,25,48), we did not quantitatively assess knowledge change among trained peer-mobilisers.

Existing community engagement strategies employed by the study clinics provided a foundation to the activities conducted under this pilot study. This study particularly benefited from working with existing mobiliser networks familiar with GBMSM- and TGW-friendly study clinics. This approach cannot be generalised to contexts with inexperienced peer mobilisers or in non-GBMSM and TGW-friendly clinics. Finally, significant time was required from the study team to deliver training, conduct weekly meetings, and provide daily supervision. As such, the frequency and length of activities are a key consideration for scalability in other contexts.

However, we believe that our study contributes to the understanding that community-engagement research is essential to addressing health inequities (49). Investing in mobilisers and their peer networks should be capitalised upon to end AIDS and reach the 95-95-95 prevention and care targets (6). The mobilisers in our study were highly motivated and committed to continuing their role beyond the study period. As new testing procedures can be unfamiliar and complex topics for mobilisers and clients, future efforts may benefit from focusing on one new concept at a time (i.e., OST or point-of-care HIV RNA testing) and dedicated training sessions and resources should be provided to facilitate these efforts. Mobilisation programmes tailored to TGW should be further explored to meet the specific needs of this populations (46,47).

## Conclusion

This study demonstrates that intensive engagement with mobilisers, through meetings and training sessions, can enhance their mobilisation skills. The continued empowerment of clients by mobilisers was essential to build trust between mobilisers and clients to ultimately motivate clients to test for HIV. Mobilisers thus play an important role in navigating their clients to access HIV testing at clinics experienced in working with LGTBI people. New concepts and testing procedures, such as AHI and point-of-care HIV-1 RNA testing, may be unfamiliar and present challenges to effective sensitization.

## Supplementary data

Supplementary materials are available in Annex 1 and Annex 2. Consisting of data provided by the authors to benefit the reader, the posted materials are not copyedited and are the sole responsibility of the authors, so questions or comments should be addressed to the corresponding author.

## Author contributions

S. P., M. D., and E. J. S. designed the study. S.P., M. D., A. K.., K. M., N. M, S. M., E. G., and E. M. E. conducted the study and collected the data. S. P., M. D., and K. M. analyzed the data. S. P. drafted the manuscript. M. D. and E. J. S. provided overall oversight for fieldwork, supervision, and manuscript writing. All authors critically reviewed and revised the manuscript and approved the final version for publication.

## Data Availability

All relevant data are within the manuscript and its Supporting Information files. Consisting of data provided by the authors to benefit the reader, the posted materials are not copyedited and are the sole responsibility of the authors, so questions or comments should be addressed to the corresponding author.

https://doi.org/10.1093%2Fofid%2Fofab219

## Acknowledgements

We would like to acknowledge all participants who took part in this study. We further acknowledge the peer mobilizers in Malindi and Mtwapa who were invaluable for mobilizing participants and their support in both this study and the parent study, and the following lesbian, gay, bisexual, transgender and intersex organizations involved in the study: Malindi Desire Initiative, AMKENI Malindi, HAPA Kenya, PEMA Kenya, GALCK, and ISHTAR MSM Health and Social Wellbeing. We would also like to acknowledge the Kenya Medical Research Institute (KEMRI)–Wellcome Trust Research Programme clinic staff who made this study possible: Abdallah Wesonga, Oscar Chirro, Joseph Nzioka, Maxwell Ong’aro, Fred Ogada, Elizabeth Wahome, Rufus Gathitu, Lucie Ikumi, Jennifer Kanungi, Riziki Rodgers, and John Omoi.

## Disclaimer

The contents are the responsibility of the study authors and do not necessarily reflect the views of the US Agency for International Development (USAID), the National Institutes of Health (NIH), the US or UK governments, the African Academy of Sciences (AAS), the New Partnership for Africa’s

Development Planning and Coordinating Agency (NEPAD), or the Wellcome Trust. This report was published with permission from the director of KEMRI.

## Financial support

This work was supported by the <u>International AIDS Vaccine Initiative</u> (IAVI) and the <u>KEMRI</u> <u>Wellcome Trust Research Programme</u> at the Centre for Geographical Medicine Research–Kilifi, supported by core funding from the <u>Wellcome Trust</u> (number 203077). This study was made possible by the generous support of the American people through <u>USAID</u>. This work was also supported in part through the <u>Sub-Saharan African Network for TB/HIV Research Excellence</u>, a DELTAS Africa Initiative (DEL-15-006). The DELTAS Africa Initiative is an independent funding scheme of the AAS Alliance for Accelerating Excellence in Science in Africa and is supported by <u>NEPAD</u> with funding from the Wellcome Trust (grant number 107752) and the <u>UK government</u>. E. J. S. received research funding from IAVI, NIH (grant number R01AI124968) and the Wellcome Trust. M. D. received funding through a PhD Scholarship from the <u>Graduate School of Amsterdam UMC–Academic Medical</u> <u>Center</u>.

## Potential conflicts of interest

All authors: No reported conflicts of interest.

---

**Annex 1: Supplementary Table 1.** Eligibility criteria of mobilised participants and index participants. AEHI, acute or early HIV infection; PrEP, pre-exposure prophylaxis; STI, sexually transmitted infection. a. Based on published behavioural PrEP eligibility score (Wahome E et al. An Empiric Risk Score to Guide PrEP Targeting Among MSM in Coastal Kenya. AIDS Behav. 2018;22(Suppl 1):35-44.) and AEHI symptom score (Sanders EJ et al. Targeted screening of at-risk adults for acute HIV-1 infection in sub-Saharan Africa. AIDS. 2015;29 Suppl 3:S221-30.)

| Study population | Eligibility criteria |
| --- | --- |
| Mobilised participants | Inclusion criteria:<br>• Age 18 years or older;<br>• Male sex assigned at birth;<br>• Reporting oral or anal sex with men in the previous six months;<br>• HIV negative or of unknown HIV status before doing the self-test;<br>• Willing to provide a blood sample;<br>• Reporting at least one behavioural factor or at least two AEHI symptoms or at least one STI symptom<sup>a</sup><br>○ Behavioural factors: in the previous seven days: any condomless sex; or in the previous three months: sex with only men, receptive anal sex or group sex;<br>○ AEHI symptoms: in the previous 14 days: fever, diarrhoea, fatigue, body aches, sore throat;<br>○ STI symptoms: in the previous 14 days: genital ulcer, genital discharge or anal discharge<br>Exclusion criteria:<br>• Well-adherent to PrEP |
| Index participants | Inclusion criteria:<br>• Age 18 years or older;<br>• Male sex assigned at birth;<br>• Reporting oral or anal sex with men in the previous six months;<br>• Being newly diagnosed with HIV, either after peer mobilisation, AEHI screening, HPN or at the HTC of one of the study clinics during the study period (April through August 2019);<br>• Willing to provide a blood sample |

**Annex 2: Supplementary Table 2.** Additional quotes.

| Quote | Respondent | Gender | Age (years) |
| --- | --- | --- | --- |
| **Experience with the training programme** |  |  |  |
| I was taught on how to deal with peers, how to talk to them and the roles of mobilizers, so I learnt a lot and now I can associate with people easily. | Mobiliser | Male | 31-35 |
| I think they [training sessions] were very helpful because we used to remind each other of things we might have forgotten because some of us were starting to maybe forget their roles and in those meeting, we help each other out. | Mobiliser | Male | 31-35 |
| **Mobilisation skills: *Networking*** |  |  |  |
| Personally, I wasn't able to talk to people one on one, but it has helped me a lot in terms of socialization and courage. Secondly, I have learnt to be discreet and conceal one's secret. | Mobiliser | TGW | 26-30 |
| Some approach me and want services... though I am not a doctor they recognize me as a doctor and I help them by taking them to [the clinic]. | Mobiliser | Male | 21-25 |
| I explained the advantages of knowing your status early and if you're not infected you will be assigned to PrEP and if you are infected you will be assigned to ART, and if you adhere to your medication your virus will be suppressed and there are no disadvantages. Either way, if you test positive or negative its for your own benefit. | Mobiliser | Male | 31-35 |
| I get my clients [peers] from my hot spot, it's where we all hustle, and they are people I know so I talk to them, and they accept being my clients. Apart from the hot spot I used to get some from social media in WhatsApp groups. | Mobiliser | TGW | 21-25 |
| **Mobilisation skills: *Client Empowerment*** |  |  |  |
| I met a peer educator at HAPA Kenya [LGBTQ organization], and he told me about KEMRI and oral self-testing. I also showed him the rashes and he empowered me to test for HIV, he gave me a lot of information and support. | Client | Male | 26-30 |
| There are some friends of mine who advised me to test. I woke up not feeling so well and decided to come to hospital then I remember what my friends had told me and decided to look for one of them who works here at the clinic. | Client | Male | NA |
| They were not willing to comply, so I used to tell them about my status and how I was. So they gained confidence from that and they decided to test and accepted to come here. | Mobiliser | TGW | 26-30 |
| What I understood from the conversation is that, it mostly touched on one's health as an adult. I could not ignore such information. | Client | Male | 26-30 |
| That's when I decided to come to KEMRI for the services, and when I came, I was served well and I confirmed my results to be truly HIV positive. He really helped me know my status. |  |  |  |
| I would talk to my peers and tell them that their health is important, and they alone can make a difference and take good care of their health. We [as mobilizers] mobilize them and it's their choice if they want to live a healthy life or not so I would encourage them to think about their health first. | Mobiliser | TGW | 29 |
| He looked at my condition and advised me to come for testing, but I was not convinced at all. In a period of two weeks he came to me again and told me it was important to come to the hospital and seek help, that's when I gave myself courage and came along with him to this place and was tested for HIV. | Client | Male | NA |
| If you hide you will not only be hiding yourself but you will be threatening your health which could be easy if you could open up to the mobiliser, because I opened up to the mobiliser, he had the courage to explain to me what is going on, which I felt supported that's why I took the step to seek for treatment through him. | Client | Male | NA |
| He told me the importance of knowing your HIV status, and if you know your HIV status you'll also get to know your partners HIV status at the same time. | Client | Male | 25 |
| **Facilitators and motivations for HIV testing** |  |  |  |
| There are some friends of mine who advised me to test. I woke up not feeling so well and decided to come to hospital then I remember what my friends had told me and decided to look for one of them who works here at the clinic. | Client | Male | NA |
| They [clients] like tell me that maybe they feel pain when they urinate and its then I start asking them questions and try to link the symptoms and inform them on the benefits of having early treatment before they become chronic and that maybe they are STIs. | Mobiliser | TGW | 26 |
| The reason why I came to get tested is that, I was not feeling well and there were some changes happening, which to me were not normal that's why I went to get tested for HIV. | Client | Male | 26 |
| On listening since I was close to the group I thought no this concerns my health, I joined them. When I listened, the information was good and I decided to join in and do a HIV test which turned positive, and we agreed to come to KEMRI clinic for further tests to confirm this status. | Client | Male | 22 |
| **Challenges with mobilisation: *Misconceptions regarding OST and AHI*** |  |  |  |
| When I received the results, I didn't believe them. I didn't trust the OST completely since I knew that if one is tested that blood must be drawn first. That's why I ignored the OST results and asked the peer mobiliser to take me with the others to [the clinic] and he agreed. | Client | Male | 22 |
| Some of my clients were used to finger prick so they were really not familiar with OST... they find it confusing but to some it was an easy option because they did not want anything to do with blood... but then you tell them if they want to know more it is best for them to do the RNA test. | Mobiliser | TGW | 25 |
| Firstly, you tell a client about AHI and the symptoms and that they are malaria-like, so clients don't see the need to test because you used 'malaria-like' and they just take painkillers and feel a little better and they don't bother about testing, so they are in denial and ignore that it might be AHI. | Mobiliser | TGW | 26-30 |
| **Challenges with mobilisation: *Logistical and financial issues*** |  |  |  |
| What I am suggesting is AHI and mobilization targets should be different because the mobilisers might lose focus and find it hard to do his job or even give an inaccurate feedback. | Mobiliser | TGW | 21-25 |
| My challenge was finding the right time for my clients [peers] because most of them are working class and they are only available during the weekends and in the evening, and they sometimes suggest that the doctors personally take those services to them. | Mobiliser | TGW | 21-25 |
| **Challenges with mobilisation: *Stigma and security concerns*** |  |  |  |
| Some of my clients were not ready to come to the centre because if they are seen accompanied by you they are afraid of being judged and questioned why they are talking to a gay... so I used to walk ahead and they would follow me. | Mobiliser | Male | 31-35 |
| In terms of harassment, Kilifi station is not pre because most people around there are homophobic and the moment you approach someone and they say you are gay you will get beaten up or even burned alive so you just have to hold back because if you even dare to be persistent and try to persuade them, they might cause you harm. | Mobiliser | TGW | 21-25 |
| My experience with trans[gender women] is that they mostly stay indoors during the day... and when people come across a trans person they will question their gender... and they might get beaten up and so... they are afraid of what will happen to them. | Mobiliser | TGW | 21-25 |
| **Recommendations for mobilisation: *Continued involvement of mobilisers*** |  |  |  |
| We should not be forgotten because we already have peers, and some are already used to us... we will have a hard time cutting off communication with clients and they will lose morale in their regular checkups and we will not be able to make follow ups because of money. | Mobiliser | Male | 18-20 |
| Mobilizers should be involved in new projects and be given priority in such matters, because it is us who have peers and recruit them and even already have established a strong connection with some, so we should be involved more on matters concerning new projects and get more training to make us more efficient with our work. | Mobiliser | TGW | 21-25 |
| What I think is we need more funds to still make follow ups on clients who did not qualify for the study so as to make them feel appreciated and maintain a close connection with them. | Mobiliser | Male | 18-20 |
| What I suggest is as this project is coming to an end we need a new project because these peers are already used to us because we some need to reach to more people who are unaware too. | Mobiliser | TGW | 21-25 |
| **Recommendations for mobilisation: *Additional training and dissemination*** |  |  |  |
| Going to the health facilities to get to know their HIV status is hard. I'm suggesting that in forums and gatherings we should be told more about HIV, the importance of frequent testing and dangers of not knowing your status in time. If people get to know more about the self-testing kits this will help people feel the ease of getting to know their HIV status and giving them the privacy they need. | Client | Male | 26-30 |
| Personally, I think we should be involved in your activities more often and it's not that whenever there is a project is when you involve us. Secondly, we need you to listen to us like the involvement in your projects, in trainings there is the questionnaire and hear out our opinions so we at least feel like we are recognized and it will give us morale. | Mobiliser | TGW | 21-25 |
| What I was suggesting is if we could get access to a video clip that shows how the machine operates so we can share the video to clients and they will have a better picture of what the RNA machine does. | Mobiliser | Male | 31-35 |
| In my opinion I think RNA test can really encourage people to test, because most of us don't like to test regularly just to confirm if we are actually negative, so this RNA test might boost testing uptake. | Client | Male | 26-30 |
| **Recommendations for mobilisation: *Tailored outreach to other key and vulnerable populations*** |  |  |  |
| What I can add on that is trans people are really something we forget to look for, so it is up to us as peer mobilizers to probe for the issues and educate them and have them over the center for their own benefits. | Mobiliser | Male | 31-35 |
| I was suggesting for road shows or campaign and so rural people can get to know about it. | Mobiliser | Male | 31-35 |
| A very low percent of trans come out in issues regarding HIV services, but if you tell them directly it is hard for them to show up. Mostly when you offer psychosocial support or such it will attract trans people and in the process of them coming, you engage them at HIV activities. | Mobiliser | Male | 21-25 |

## Footnotes

1 GBMSM and TGW will be used as an inclusive term throughout this paper, including both men who have sex with men (MSM) and transgender women. MSM denote sexual behaviour, including men who do not identify as LGBTI and transgender women, for whom the term MSM erases their gender identify.

## Notes

### Competing Interest Statement

The authors have declared no competing interest.

### Funding Statement

This work was supported by the International AIDS Vaccine Initiative (IAVI) and the KEMRI Wellcome Trust Research Programme at the Centre for Geographical Medicine Research Kilifi, supported by core funding from the Wellcome Trust (number 203077). This study was made possible by the generous support of the American people through USAID. This work was also supported in part through the Sub Saharan African Network for TB/HIV Research Excellence, a DELTAS Africa Initiative (DEL 15 006). The DELTAS Africa Initiative is an independent funding scheme of the AAS Alliance for Accelerating Excellence in Science in Africa and is supported by NEPAD with funding from the Wellcome Trust (grant number 107752) and the UK government. E. J. S. received research funding from IAVI, NIH (grant number R01AI124968) and the Wellcome Trust. M. D. received funding through a PhD Scholarship from the Graduate School of Amsterdam UMC Academic Medical Center

### Author Declarations

The KEMRI Scientific Ethical Review Unit of the KEMRI Wellcome Trust Research Programme gave ethical approval for this work (Study code 135/3747).

